# Towards validation of clinical measures to discriminate between nociceptive, neuropathic and nociplastic pain: Cluster analysis of a cohort with chronic musculoskeletal pain

**DOI:** 10.1101/2024.08.13.24311924

**Authors:** Paul W Hodges, Raimundo Sanchez, Shane Pritchard, Adam Turnbull, Andrew Hahne, Jon Ford

## Abstract

The International Association for the Study of Pain defines three pain types presumed to involve different mechanisms - nociceptive, neuropathic and nociplastic. Based on the hypothesis that these pain types should guide matching of patients with treatments, work has been undertaken to identify features to discriminate between them for clinical use. This study aimed to evaluate the validity of these features to discriminate between pain types. Subjective and physical features were evaluated in a cohort of 350 individuals with chronic musculoskeletal pain attending a chronic pain management program. Analysis tested the hypothesis that, *if* the features nominated for each pain type represent 3 different groups, *then* (i) cluster analysis should identify 3 main clusters of patients, (ii) these clusters should align with the pain type allocated by an experienced clinician, (iii) patients within a cluster should have high expression of the candidate features proposed to assist identification of that pain type. Supervised machine learning interrogated features with the greatest and least importance for discrimination; and probabilistic analysis probed the potential for coexistence of multiple pain types. Results confirmed that data could be best explained by 3 clusters, clusters were characterized by *a priori* specified features, and agreed with the designation of the experienced clinical with 82% accuracy. Supervised analysis highlighted features that contributed most and least to the classification of pain type and probabilistic analysis reinforced the presence of mixed pain types. These findings support the foundation for further refinement of a clinical tool to discriminate between pain types.

## Introduction

Chronic pain contributes to 71% of disability worldwide[9]. Back pain alone costs the US economy >$100 billion annually[3], the most of any condition. Thousands of randomized controlled trials show modest effects[11]. One size does not fit all, yet >90% of back pain is labelled “non-specific” of unknown cause[10] without evidence-based rationale to guide treatment. Trial-and-error application of treatment is wasteful, inefficient, and rejected by those with chronic pain[4]. High variability in individual responses could be improved by predictive biomarkers that guide treatment allocation. Unfortunately, attempts to target treatments have had limited success[33]. Identification of the predominant mechanism that explains an individual’s pain could help guide treatment[27].

Pain involves an array of inputs and outputs, and diverse biological and neural mechanisms, influenced by factors including emotions and cognitions[8]. Although activation of nociception by actual or threatened tissue damage is one input, many other inputs and mechanisms interplay. These differ between individuals[37]. The International Association for the Study of Pain (IASP) defines 3 pain types that are hypothesized to relate to different underlying mechanisms[27], which should require different treatments. These are Nociceptive pain – arising from actual or threatened damage to non-neural tissue, due to activation of nociceptors; Neuropathic pain – caused by lesion or disease of the somatosensory nervous system; and Nociplastic pain –arising from altered nociception despite no clear evidence of actual or threatened tissue damage causing activation of nociceptors or evidence for disease or lesion of the somatosensory system[24]. Biomarker signatures for pain mechanisms have potential to predict response to targeted treatments for chronic pain.

There is no “gold standard” to discriminate between types. Instead, methods have been proposed using multi-modal data (e.g., quantitative sensory testing (QST), pain qualities, questionnaires) to identify features suggesting predominance of one mechanism. Progress has been compromised by diversity of opinion regarding *how* pain types might be identified[16,30] and *if* this is possible[35]. Recent systematic reviews[27,28] and expert consensus[29] have identified and refined candidate measures that could aid in the discrimination between the pain types, and other work has defined clinical criteria for identification of a single type (e.g., nociplastic[15,23]; neuropathic[5]). In the absence of a gold standard tool to identify mechanism, validation has been difficult.

One alternative to evaluate the validity of the discrimination between the IASP pain types is to collect data for a cohort of individuals with diverse presentation of chronic pain, and test the hypothesis that, *if* the features nominated for each pain type represent 3 different groups, *then* (i) cluster analysis should identify 3 main clusters of patients, (ii) these clusters should align with the pain type allocated by an experienced clinician, (iii) patients within a cluster should have high expression of the candidate features that are proposed for that pain type. This study aimed to test these hypotheses using a cohort of individuals with chronic musculoskeletal pain attending a pain management clinic, and then to (i) interrogate which features had the greatest and least importance for the discrimination; and (ii) to probe the potential for coexistence of multiple pain types.

## Methods

### Study design

This study evaluated a cohort of individuals with diverse presentations of chronic pain using the highest ranked features allocated by expert consensus[29] to aid discrimination between the IASP pain types. For this study all features were assessed with simple subjective and physical examinations that were feasible in a clinical setting without specialised equipment. All items were assessed for all participants (with some limited exceptions). The natural tendency for the data to cluster in to three groups was evaluated first. If three clusters were present, we then interrogated the relationship with the predominant pain type allocated based on review and scoring of the examination results by an independent experienced clinician. Because the data assessed as “typical features” of each pain type *and* the classification of the participant’s predominant pain type use the same data, it might be argued the analysis step is biased to confirm the hypothesis. This outcome cannot be assumed because the clustering algorithms (unsupervised machine learning models) do not use the dependent variable (pain type) for modelling or prediction, that is, neither the score-derived nor examiner-derived pain type was used in generation of clusters. Second, we employed supervised machine learning to understand the relative importance of test items to the pain-type clusters. Third, we considered probabilistic models to provide insight into the coexistence of multiple pain types, which remains hidden in deterministic analysis.

### Participants

Participants were 350 individuals with chronic pain who presented for assessment at a chronic pain management program. They were included if they had a primary complaint of chronic musculoskeletal pain (pain of >3 months duration), were referred to attend a multidisciplinary pain management program, were aged between 18 and 75 years, and had sufficient fluency in English to complete the assessment. Participants were excluded if they had red flags that would be unsuitable for pain management (including active cancer, signs of cauda equina syndrome based on bladder or bowel disturbance, risk of spinal fracture, signs of potential infection, foot drop that my cause tripping, spondyloarthropathy). The Human Research Ethics committee of LaTrobe University provided ethical approval for the collection and analysis of the data. Participants were asked to provide informed consent for analysis of their de-identified data when attending their initial assessment. Characteristics of the participant group were recorded using the electronic Persistent Pain Outcomes Collaboration (ePPOC) standardised assessment data set[34].

### Measures

Features were assessed as part of the clinical examination. Items were selected from the highest ranked items from the recent expert consensus study[29], and refined using the clinical criteria reported to identify nociplastic[15,23] and neuropathic[5] pain. The full list of features considered for each pain type are presented in Supplementary Data 1-3. From the candidate items, a list of subjective and physical examination items was generated that was possible to apply in a clinical setting as subjective or physical tests. Table 1 presents the final list of features. The 31 items included 11 for nociceptive pain, 8 for neuropathic pain and 12 for nociplastic pain. For nociplastic pain, physical item 3 was only completed if item 2 was negative. All items were scored on a scale of 0-100, anchored with “not present” at 0 and strongly present at “100”. Although all items were used as independent items in the analysis, we also calculated the average score for the questions for each pain type separately. The dominant pain type was allocated based on that with the highest average score (Score-derived pain type). If a participant presented with multiple pain locations, only the “main pain location” was scored. An experienced clinician independently reviewed the patient’s medical record, including the response to the pain type questions and assigned a likely predominant pain type (Examiner-derived pain type).

**Table 1.** Variables considered in the study.

| **Mechanism** | **Category** | **#** | **Item** |
| --- | --- | --- | --- |
| **NOCICEPTIVE**<br>**CRITERIA** | **Subjective**<br>**examination** | **1** | Localised distribution of pain |
|  |  | **2** | Proportional and direct relationship of pain response with aggravating factors |
|  |  | **3** | Pain provocation by specific postures and/or movements consistent with a nociceptive clinical pattern |
|  |  | **4** | Findings from imaging of body regions of potential relevance to the pain experience |
|  |  | **5** | Below cut-off (< 0) on Neuropathic Pain Questionnaire |
|  | **Physical**<br>**examination** | **1** | Signs of inflammation or a positive Clinical Inflammation Score |
|  |  | **2** | Proportional and direct relationship of pain response with physical examination |
|  |  | **3** | Pain provocation by testing of specific postures and/or movements consistent with a nociceptive clinical pattern |
|  |  | **4** | Pain provocation by special tests consistent with a nociceptive clinical pattern |
|  |  | **5** | No generalised hypersensitivity |
|  |  | **6** | Absence of autonomic symptoms and/or signs (from CRPS features) |
| **NEUROPATHIC**<br>**CRITERIA** | **Subjective**<br>**examination** | **1** | Dermatomal or peripheral nerve distribution of pain |
|  |  | **2** | Dysesthesia (eg electric shock-like, lightning, paraesthesia) |
|  |  | **3** | Evidence of lesion or disease of the nervous system |
|  |  | **4** | Hypoaesthesia in a neuroanatomically plausible distribution |
|  |  | **5** | Provoked by movements that load or compress neural tissue |
|  |  | **6** | Above cut-off (>0) on Neuropathic Pain Questionnaire |
|  | **Physical**<br>**examination** | **1** | Sensory deficits in a neuroanatomically plausible distribution |
|  |  | **2** | Positive findings on nerve pain provocation testing |
| **NOCIPLASTIC**<br>**CRITERIA** | **Subjective**<br>**examination** | **1** | Sensitivity to light touch or pin prick |
|  |  | **2** | Sensitivity to deep pressure |
|  |  | **3** | Sensitivity to heat or cold |
|  | **Presence of**<br>**comorbidities** | **1** | Increased sensitivity to sound and/or light and/or odours |
|  |  | **2** | Sleep disturbance with frequent nocturnal awakenings |
|  |  | **3** | Fatigue |
|  |  | **4** | Cognitive problems such as difficulty to focus attention, memory disturbances, etc |
|  | **Physical**<br>**examination** | **1** | Dynamic mechanical allodynia to light touch |
|  |  | **2** | Static mechanical sensitivity to deep pressure |
|  |  | **3** | Heat or cold sensitivity (only test if above are negative) |
|  |  | **4** | Painful after-sensations reported following the assessment of any of the above |
|  | **Other**<br>**Criteria** | **1** | Regional distribution |

### Data analysis

#### Data preparation

Quality of data was assessed in several ways depending on data type. Independent variables: The dataset contained responses on a scale from 0 to 100. Missing data were analysed through a graphical distribution analysis of the percentage of missing data. We assumed that missing metrics were ignored by the examiner because they were not necessary to allocate a pain type to the participant (e.g., *nociplastic physical 3* was not performed if features 1 or 2 were positive). For this reason, missing values were imputed with zeros to avoid losing sample size.

Dependent variables: Dominant pain mechanisms were analysed using a distribution plot to consider the generalization ability of the model, and to consider class balance or imbalance for selection and configuration of the clustering algorithm. This was undertaken separately for pain types derived from pain scores, and that diagnosed by an examiner. Unambiguous diagnosis was present in 80% of cases.

#### Pre-clustering analysis using descriptive statistics and correlations

A pre-clustering analysis was performed to ensure the independent variables contained sufficient information to distinguish between the three types of pain, by way of validating the potential to consider a clustering approach. First, a correlation analysis of the independent variables was performed using Pearson’s correlation, visualised through a correlation plot, and arranged by nociceptive, nociplastic, and neuropathic criteria. To support the clustering approach, a high correlation within categories and low correlation between categories would be expected.

Second, a heatmap was created, ordering data by pain type obtained from scores, visualising the “intensity” of responses to corresponding criteria to identify patterns. To support the clustering approach, patients within a cluster should have high expression of relevant features.

Third, before clustering we also computed the Hopkins statistic[17], to measure the natural cluster tendency in the data. Values close to 1 indicate that the data are highly clustered, and close to 0.5 indicates that the data are randomly distributed. Values below 0.75 suggest that the clusters are sparse and with fuzzy borders between them. For this analysis, data were reduced using principal components analysis for visualization purposes as it allows projecting n-dimensional data into 2-dimensions. The clustering tendency was computed with the original features. The clustering tendency was presented using a scatter plot with the first and second principal components on the x- and y-axes, respectively. A 2D density plot was included to visualise the concentration of pain types.

#### Consistency of clusters with pain type classification

To determine whether analysis would identify three clusters, and if so, whether these were aligned to the three pain types we analysed the data ins several ways. First, the number of clusters was evaluated, considering clusters between two and ten. Three common clustering methods were assessed: k-means, Gaussian mixture models (GMM), and Hierarchical clustering[26]. The silhouette coefficient was calculated for each scenario. This coefficient provides an indication of how well each data point was clustered, with positive values indicating better classification[25]. We determined the optimal number of clusters as that which maximized the silhouette coefficient. To support our hypothesis that the clinical features relate to three pain types, the optimal number of clusters would be three.

Second, using the three clusters, the consistency of each clustering method was evaluated by comparing each point’s clustering against the pain type derived from score-derived and examiner-derived pain types. Each of the six resulting comparisons was evaluated using the True Positive Rate (TPR), which accounts for the percentage of cases that were correctly classified according to the average score (score-derived classification) and the experienced examiner (examiner-derived classification). This value is from 0 to 100%.

Third, for hierarchical clustering (which provided the second highest TPR, see results, but enables investigation of the internal structure), a dendrogram was generated to graphically analyse the hierarchical structure, showing the model’s clusters and allocation to predominant pain type based on highest scores. Within this approach the Euclidean distance and averaged linkage grouping criteria were used to allocate participants to each cluster.

#### Prediction of pain type based on pain features using supervised machine learning and evaluation of relative contribution of test items to discrimination

If the unsupervised analysis confirmed the presence of three clusters, we planned to use supervised learning to explore the possibility of extracting relevant information to understand the relative value of individual test items to the classification. Because of the small sample size we applied a logistic regression model, which has fewer parameters than other models, allows the prediction of pain type based on the independent variables, and interprets the importance of the features. This approach was applied to the classification of the predominant pain type deduced from the highest scores and that allocated by the independent examiner. We used 80% of the data for model training. The coefficients of both models were evaluated, analysing the sign of these coefficients to determine each variable’s contribution to the prediction. The accuracy of both models was also evaluated using the TPR, ensuring consistency with the previous clustering comparison.

#### Probabilistic predictions of pain mechanisms to investigate coexistence of pain types

Finally, a probabilistic analysis was conducted to explore the uncertainty in model predictions, both supervised and unsupervised. GMM and logistic regression were used to output probabilistic classifications, and a probabilistic variation of k-means called c-means was also used to generate cluster membership likelihoods. The likelihood of the dominant pain type for each model is presented as boxplot. Values that are concentrated around 100% show the model is more certain about the prediction, whereas sparse values show higher uncertainty. Each comparison is presented with the Fuzzy Partition Coefficient (FPC), which evaluates the degree of uncertainty in the predictions, with values closer to 100% indicating well-defined and distinct clusters (i.e., if all pain types were mutually exclusive, the FPC would be 100% for all pain types).

## Results

### Participants

Data were available for 350 participants presenting for pain management with a primary complaint of chronic musculoskeletal pain. Participants (62.7% female; 37.3% male) had an average (standard deviation) age, height and weight of 46(12) years, 167(11) cm and 84.2(20.9) kg, respectively. Regarding the duration of pain, 1.7% of individuals had experienced pain for less than 3 months (at the time of completion of the questionnaire, but >3 months at the time of testing), 22.3% for 3 to 12 months, 33.9% for 12 months to 2 years, 29.8% for 2 to 5 years, and 12.4% for more than 5 years. When asked to describe their pain, 0.4% reported that it was rarely present, 4.7% said it was occasionally present, 8.5% described it as often present, 75.3% stated it was always present with varying intensity, and 11.1% reported it was always present with consistent intensity. The main sites of pain reported were the low back (28.3%), shoulder (22.2%), and neck (19.1%), with other less common sites including the mid back (5.7%), knee (4.8%), foot (3.9%), hip (3.5%), wrist (2.6%), hand (2.6%), and other areas (7.4%). The average total Pain Disability Questionnaire Score was 104 (27). From the possible data 265 data points contain at least one variable with missing values.

### Data preparation

Figure 1 shows the distribution of missing values which indicates that most metrics have <15% of missing data. As expected, metrics such as “*nociplastic subjective 3*” (See Table 1 for item definitions) and “*nociplastic physical 3*” have the highest percentage of missing values, exceeding 30%. Pain type derived by the examiner was unavailable for 20% of participants. In this case the experienced examiner had identified the pain type as ambiguous.

**Figure 1.**
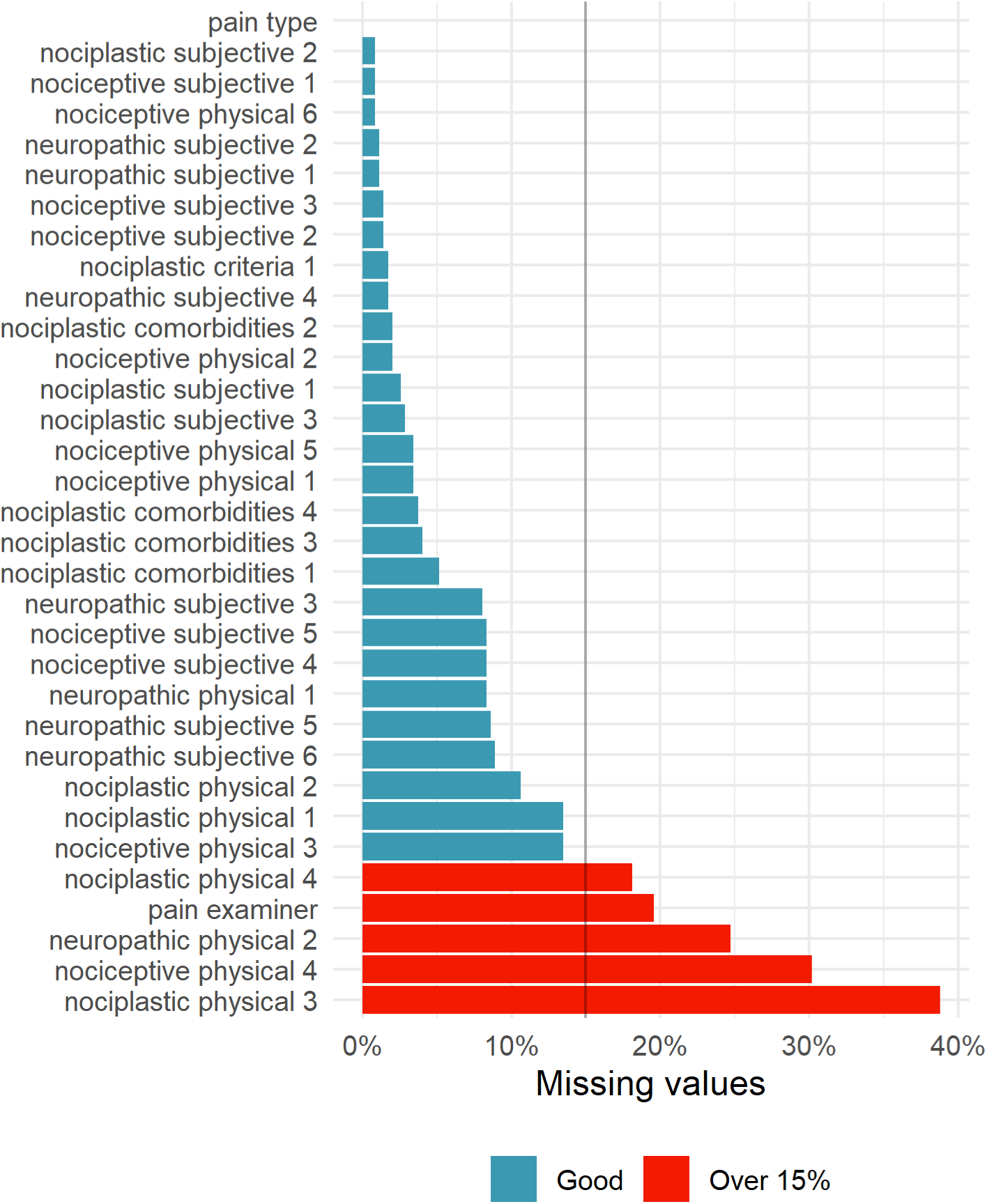
Percentage of missing values for the different variables of the study. Blue bars represent metrics with less than 15% missing data, whereas red bars represent metrics with more than 15% missing data.

The available sample is unbalanced for the different pain types (Figure 2). According to the pain type (both score-derived or examiner-derived), most cases were classified as nociplastic, followed by nociceptive and neuropathic with the least cases. In the case of the examiner’s unambiguous classification, all nociceptive participants are available, but availability of classification of the other pain types was reduced by diagnostic uncertainty.

**Figure 2.**
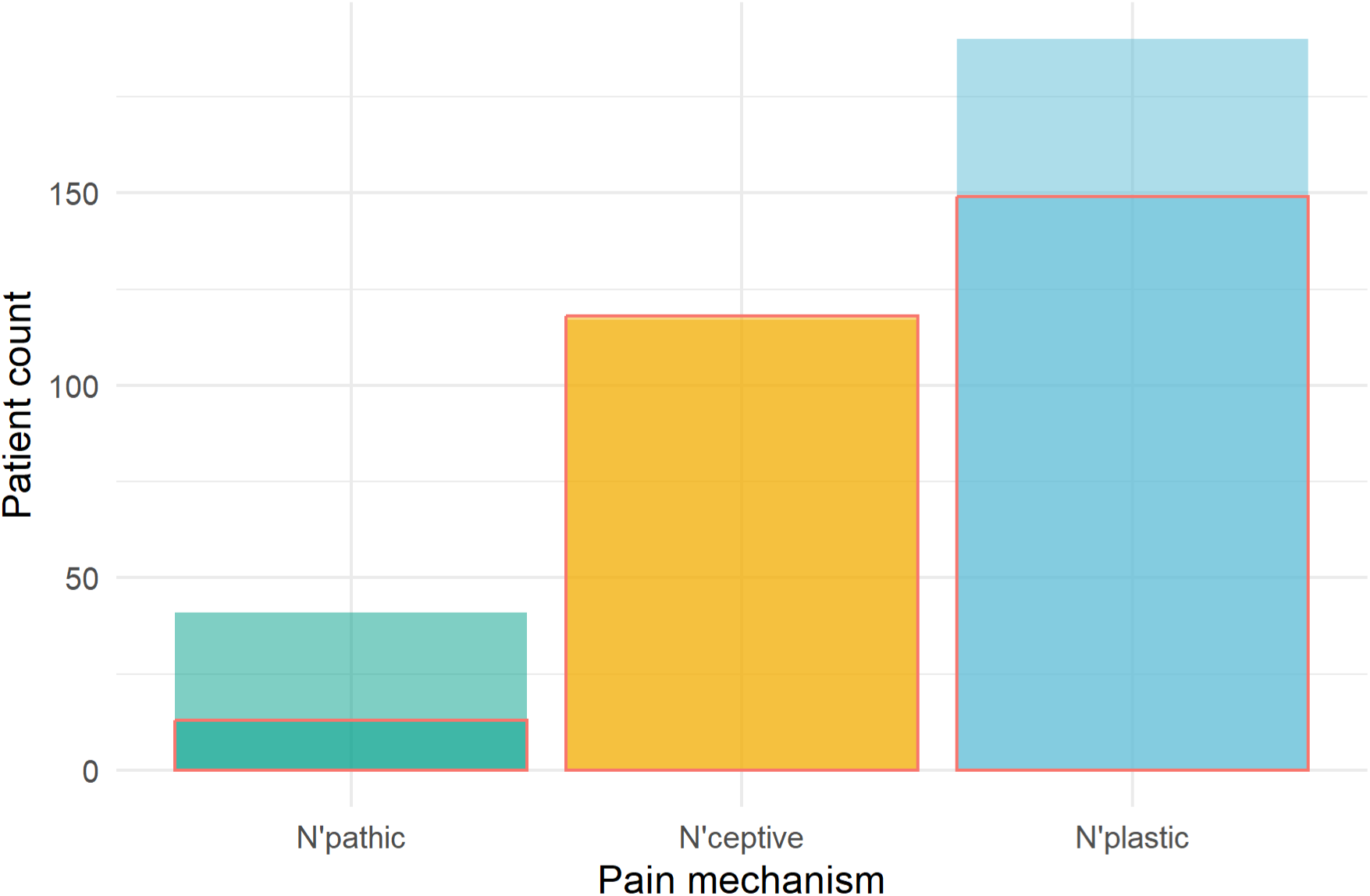
Distribution of participants across three different pain types. The y-axis represents the patient count for each pain mechanism, and the red outline box indicates the examiner’s available classification for each pain type. In the cases where the experienced examiner did not allocate a classification it was indicated as ambiguous, this uncertainty was higher in participants with higher neuropathic scores. This might bias the results using examiner-derived pain types, particularly for underrepresented groups such as those with neuropathic pain.

### Pre-clustering analysis using descriptive statistics and correlations

As we required to proceed to the clustering step, the correlation among variables was higher and positive within the criteria for each pain type, than between pain types (Figure 3; high correlation expected within each red box). This finding demonstrates strong internal consistency. The highest correlation (R=0.82) was between *nociceptive subjective items 2* and *3*, which both relate to the relation of pain to aggravating factors. Subjective and physical features of nociceptive that address similar constructs are highly correlated. Negative relations can be seen between features of different pain types, such as nociceptive and nociplastic, where negative values present in almost all comparisons. *Nociceptive subjective 5* and *neuropathic subjective 6* have the strongest negative correlation, which is expected as these items relate to low or high scores, respectively, on the same neuropathic questionnaire. This neuropathic questionnaire was modestly correlated with nociplastic features, suggesting its items reflect both pain types.

**Figure 3.**
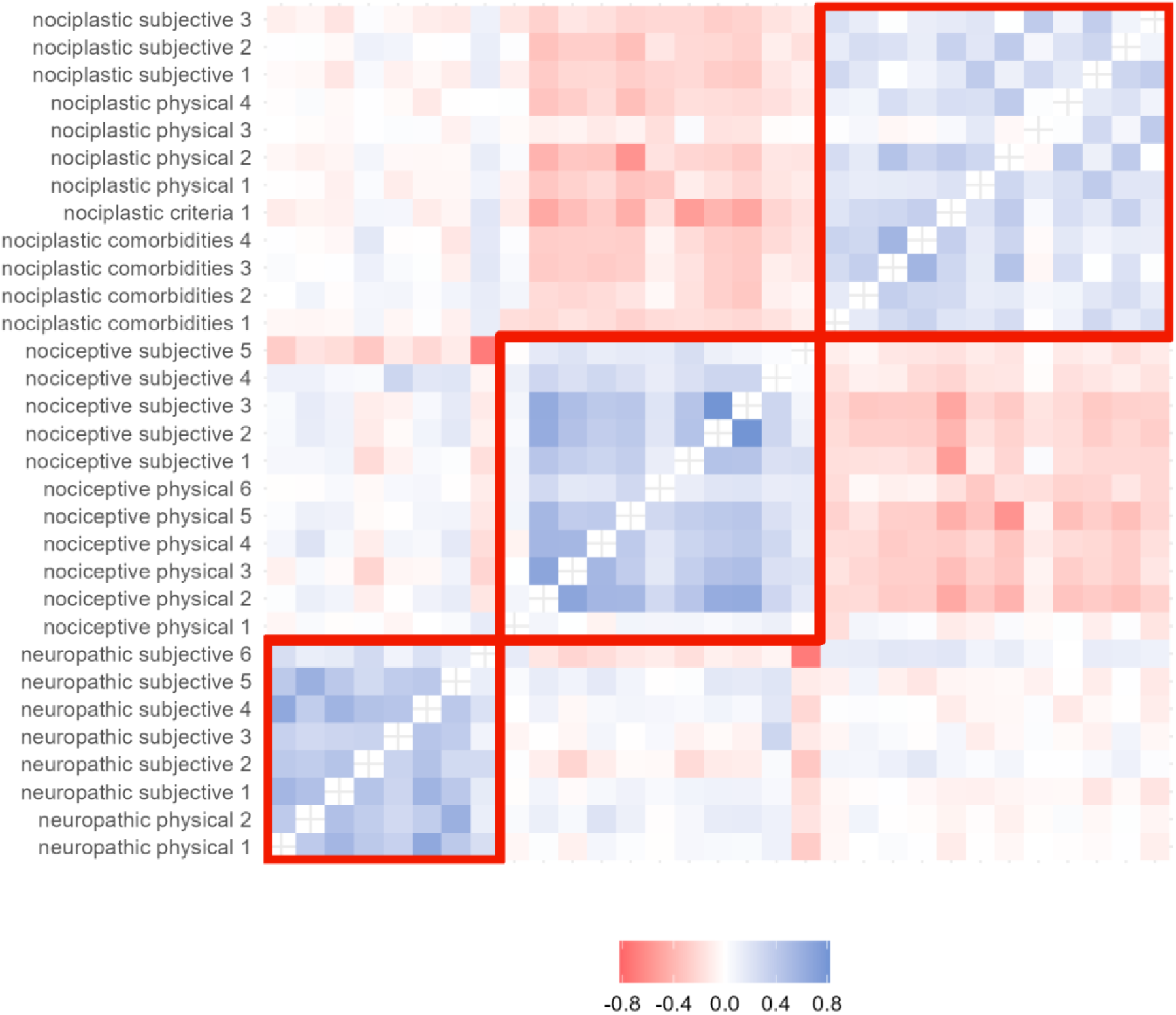
Correlation plot between the study independent variables. Each cell represents the Pearson correlation coefficient between pairs of variables, with the colour indicating the strength and direction of the correlation. Red boxes group specific questions for each pain mechanism.

The pattern of expression of each feature across participants is shown as a heat map in Figure 4. As expected, the density of blue, determined by the intensity of expression of each feature is high, with the red boxes that are bounded on the x-axis arranged by pain type classified according to the scoring, and on the y-axis by the features for each pain type. In some cases (e.g., *nociceptive physical 6*, or *nociplastic comorbidities 1*) there is stable expression across all pain types, which suggests limited value in discrimination between the pain types. Note *nociplastic physical 3* is commonly scored as 0 because it is only tested if *nociplastic physical 2* is negative.

**Figure 4.**
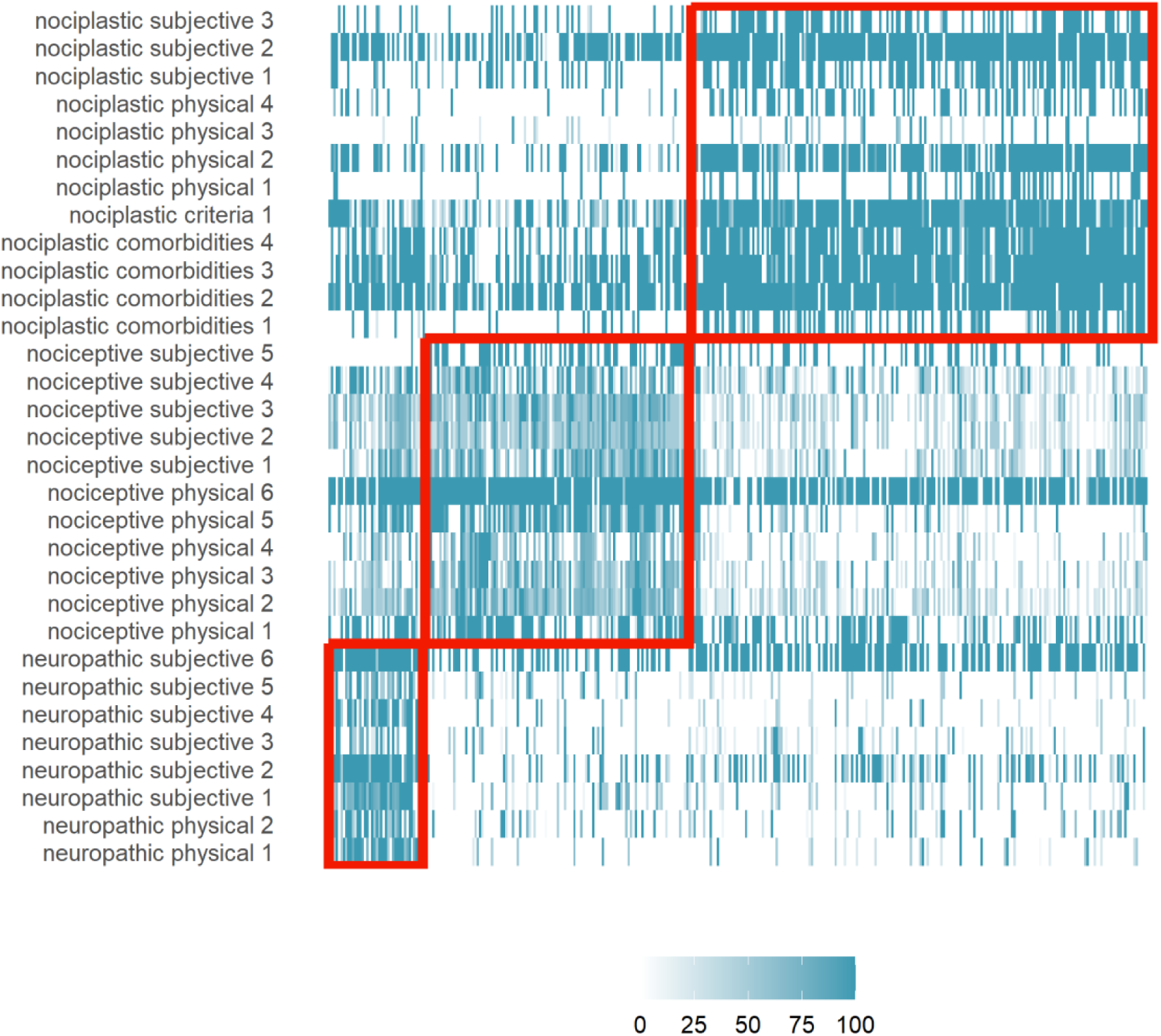
Heatmap of all the independent variables. Each cell represents the value of the participants answers to a given question, with the participants arranged by dominant pain type allocated by the largest score. Red boxes group specific questions for each pain mechanism with participants with corresponding dominant pain type.

When analysing the clustering tendency of the data, the Hopkins statistic was 60%. This indicates some degree of natural clustering is present in the data, but we should not expect perfectly defined clusters. The data distribution presented in Figure 5 shows that the density curves for pain types overlap between each other, suggesting that many points may be classified with more than one pain type. Consideration of the weightings for each of the feature in the principal components demonstrates that principal component 2 distinguishes neuropathic from the other 2 and, as expected, is weighted by neuropathic features. Further, nociceptive and nociplastic are distinguished but principal component 2, which is weighted by features of these pain types.

**Figure 5.**
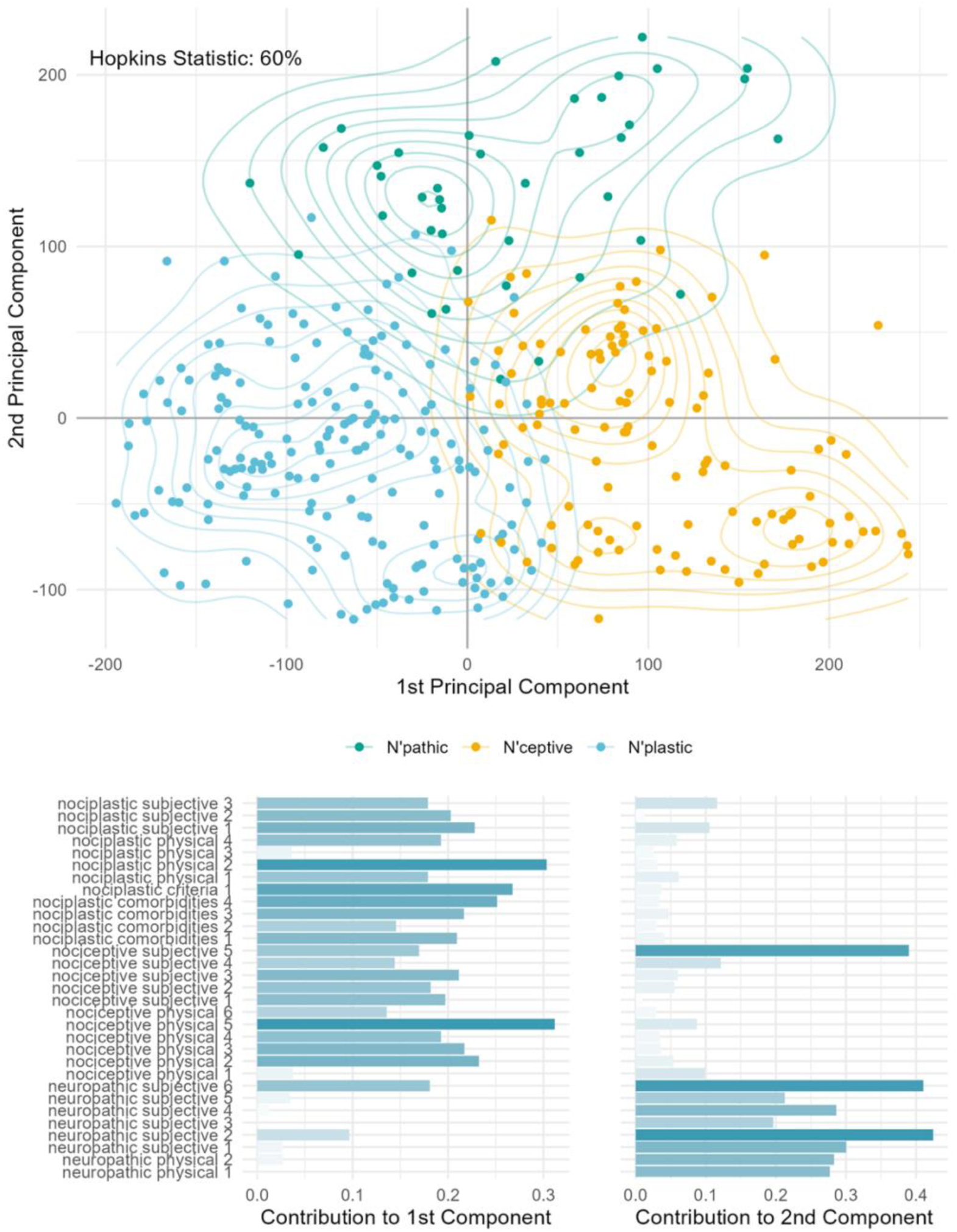
Cluster tendency of the independent variables. The upper panel demonstrates the Hopkins Statistic. The x- and y -axis represent the 1^st^ and 2^nd^ principal component of the data. For each pain type, a density map is depicted to illustrate the differentiation of pain type’s distribution. The bottom panel shows the features that contribute to the 2 principal components. Note that *nociceptive subjective 5* is a low score on the neuropathic questionnaire.

### Assessment of consistency of classification with predominant pain type

When testing the clustering algorithms, the optimal number of clusters was analysed as shown in Figure 6. The three clustering models minimize the share of negative silhouette at three clusters. At this point the overall silhouette is maximized for hierarchical approach, and for GMM and k-means is only beaten by the silhouette of using two clusters. Combining both metrics would suggest using three clusters.

**Figure 6.**
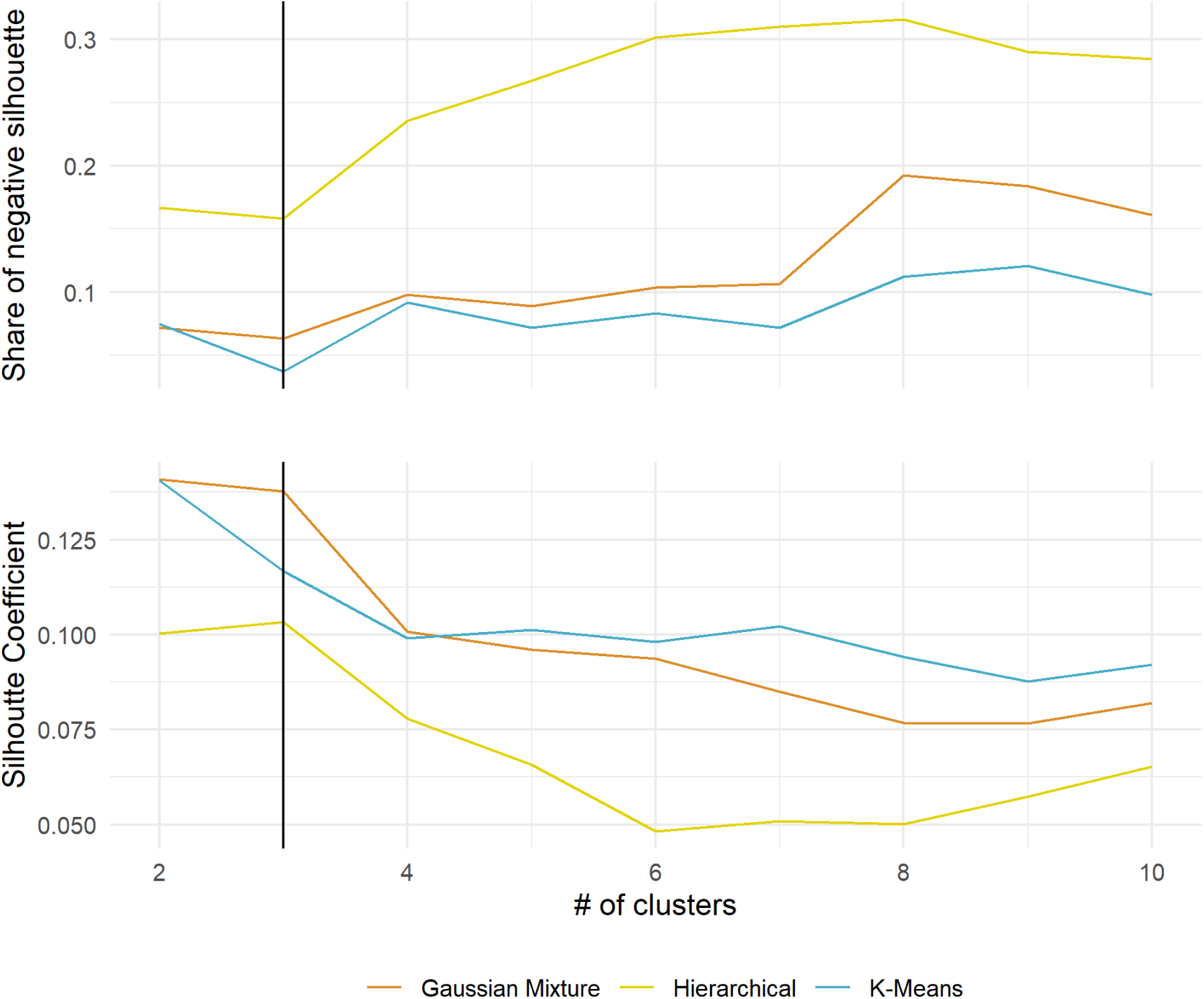
Cluster Silhouette Analysis for different number of clusters. Silhouette indicates how well each data point was clustered, so it should be maximized. Three clustering methods were tested, varying the number of clusters between 2 and 10. Low values for the share of negative silhouette also serves as an overall metric of proper clustering.

When predicting pain type using clustering methods, results differ depending on which version of the ground truth (score-derived or examiner-derived pain type) was used, as shown in Figure 7. For interpretation of this analysis, the pain type that was most likely to be represented by each cluster was identified by inspection of the heat map which showed the features that were commonly expressed in each cluster. As shown for the hierarchical cluster method in Figure 8, the largest cluster was dominated by high expression of features of nociplastic pain, and so on. When prediction accuracy was analysed using both versions, the best model was the GMM. The examiner’s diagnosis achieved a TPR of 82%, and the score derived pain type achieved a TPR of 89%.

**Figure 7.**
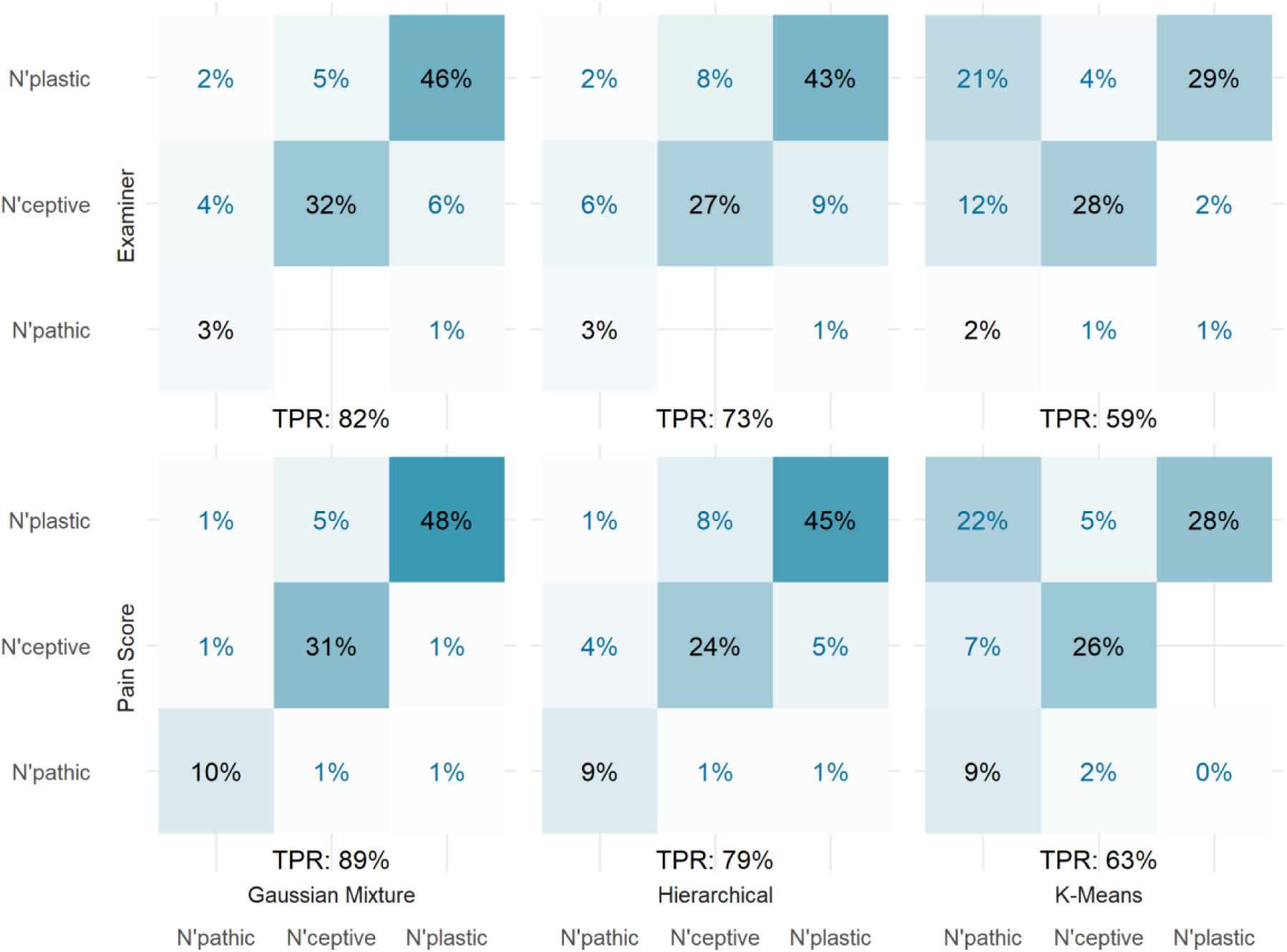
Clustering prediction accuracy. Each of the 3 clustering algorithms were tested against the dominant pain type as computed with the score, and against the pain defined by the examiner. Each comparison shows the percentage of predictions from one pain type that were classified as any pain type. The diagonal represents the correct predictions, and the sum of the diagonal is the True Positive Rate (TPR).

**Figure 8.**
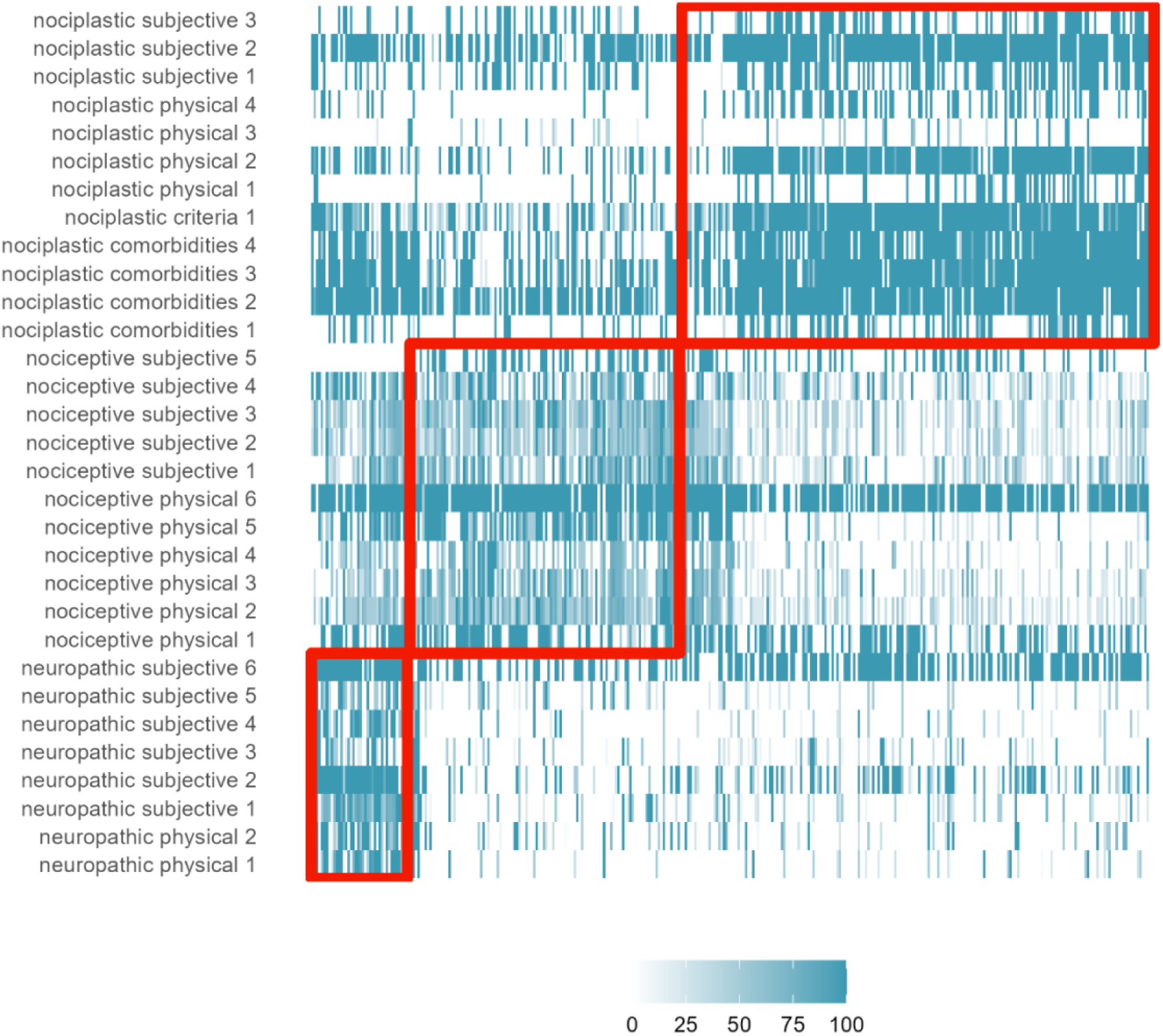
Heatmap of all the independent variables for the clustered data. Each cell represents the value of the participants answers to a given question, with the participants arranged by the three clusters. Red boxes are bounded by the allocation of the participants to a cluster in the x-axis, and the pain type specific questions on the y axis.

The hierarchical method achieved close to 80% TPR, and we can observe the structure of this model’s predictions in a tree structure (i.e., dendrogram) shown in Figure 9. It shows that the nociplastic predictions are the most consistent.

**Figure 9.**
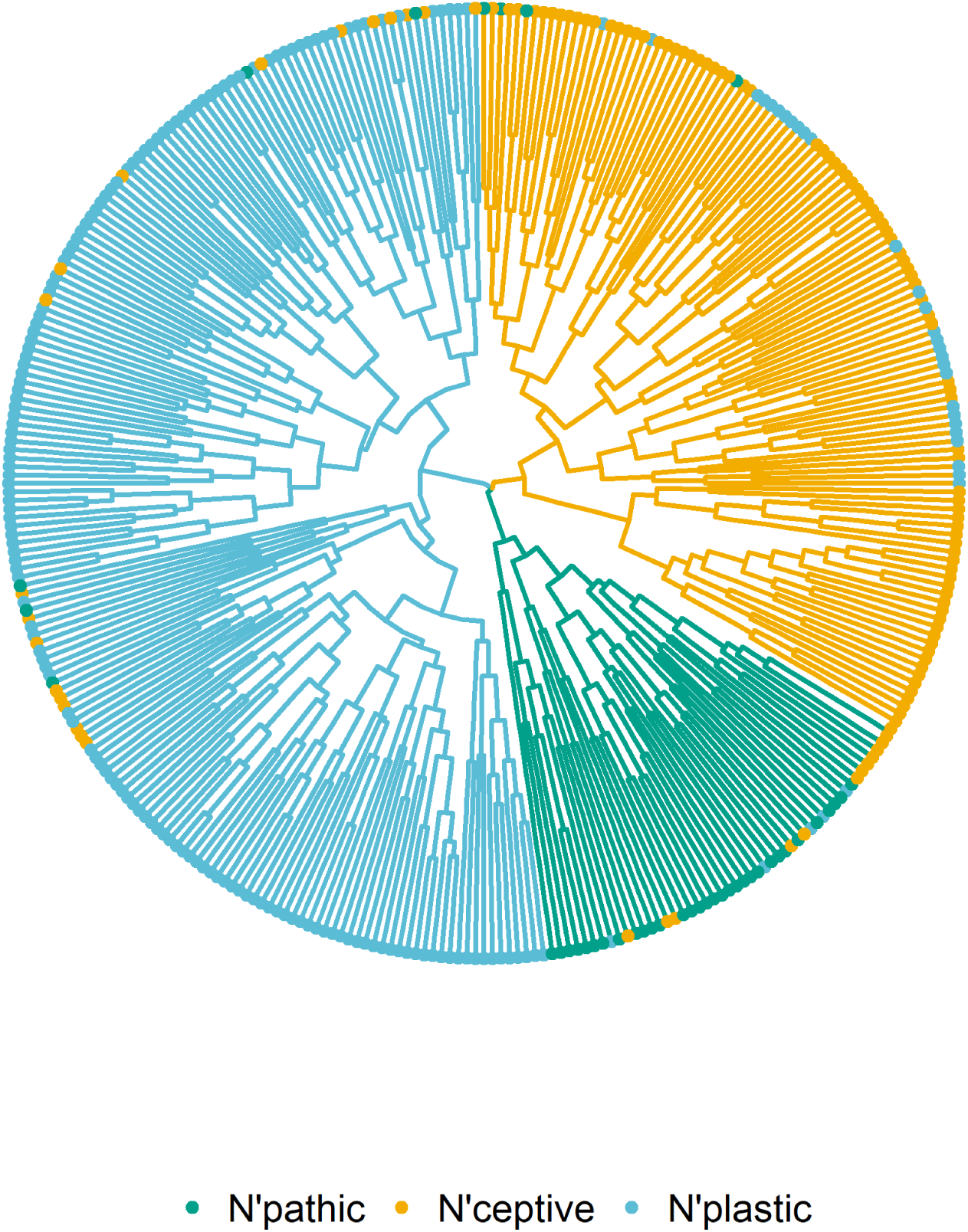
Cluster dendrogram. The tree structure represents the closeness between data points, where they merge iteratively until only one cluster is formed. The distance to the centre represents the difference between the merging branches. The colour of the branches represents the predominant pain type as defined by the model, and the colour of the points at the end of each branch represents predominant pain type defined by the pain score. Note the high degree of consistency

### Prediction of pain type based on pain features using supervised machine learning

The sign of coefficients from application of supervised machine learning using logistic regression models, were in the expected direction with positive values for features of corresponding pain type (Figure 10), and negative values for other pain types. The coefficient magnitudes were greater for the pain derived score model than for the examiner. This was expected as the score was directly derived from these measures. No features had significant positive magnitudes for more than one pain type. Several features show non-significant values for the three predicted pain types, which means they contributed little to the prediction of pain type (Table 2). Features such as *nociplastic comorbidities 2*, *neuropathic subjective 6* and *nociceptive physical 2, physical 6* and *subjective 3* contributed little to the prediction and were redundant.

**Figure 10.**
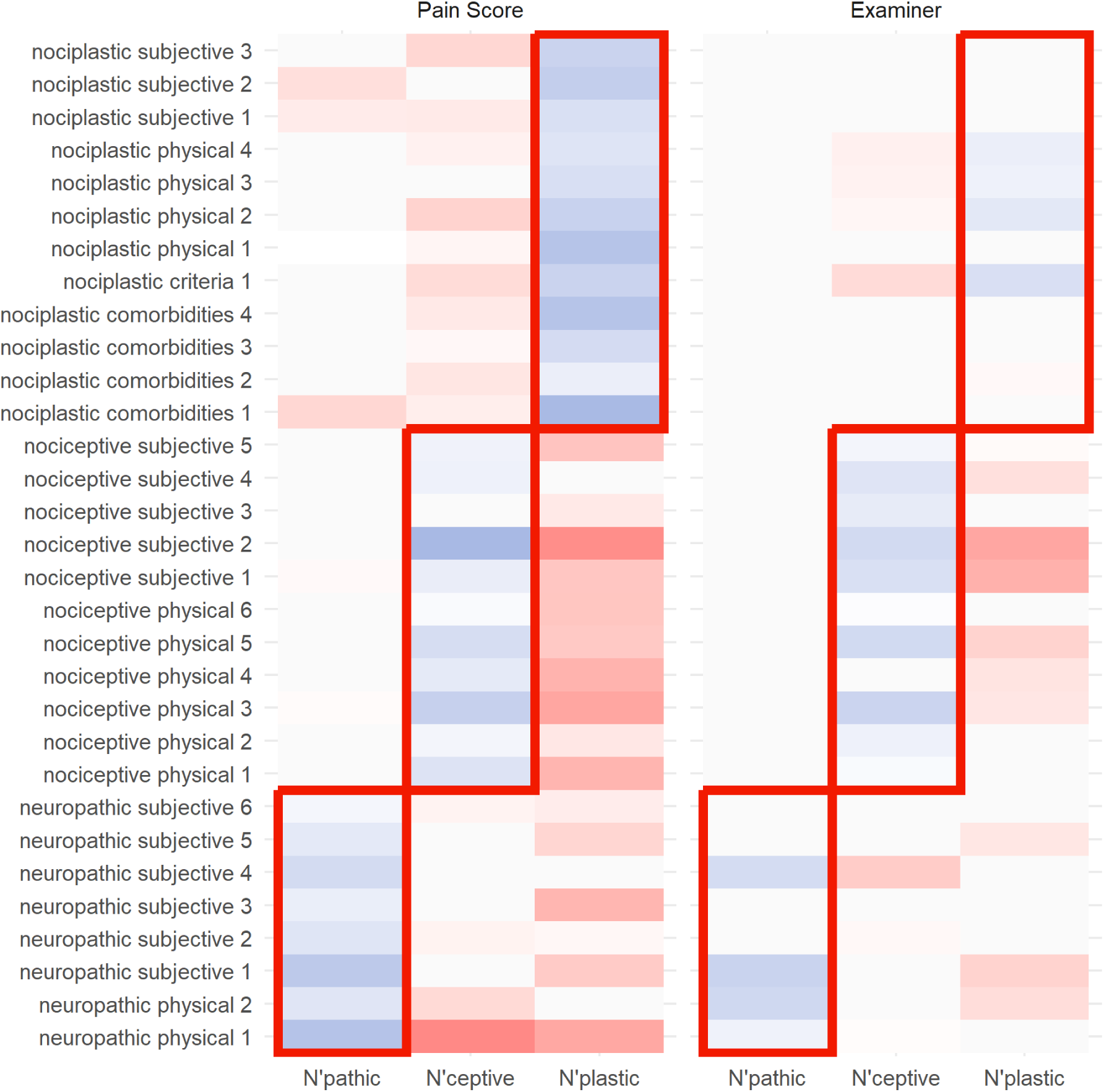
Coefficient heatmaps from logistic regression analysis for score-derived and examiner-derived pain types. The heatmap displays coefficients for each feature based on the observed pain type. Blue indicates a positive relationship, whereas red indicates a negative relationship with the pain mechanism categories. The outlined boxes on both heatmaps highlight the grouped coefficients for each pain mechanism.

**Table 2.**
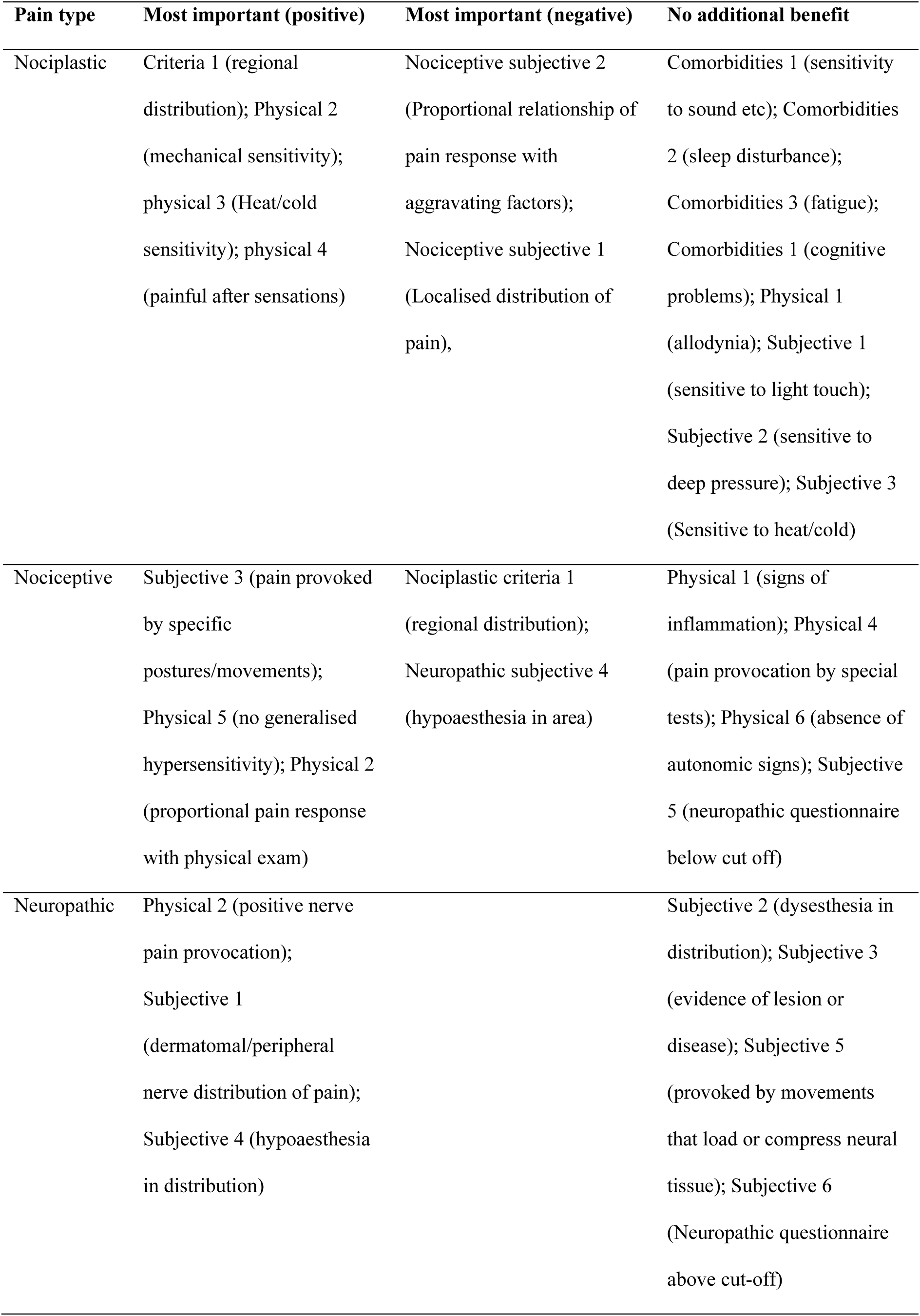
Most and least important features to support classification of pain type.

After performing predictions, the supervised classification models were evaluated with the TPR, and can be compared with previous results from cluster analysis. Figure 11 shows that the model for the score derived pain type achieves a TPR of 94%, whereas the model for the examiners pain type achieves a TPR of 87%.

**Figure 11.**
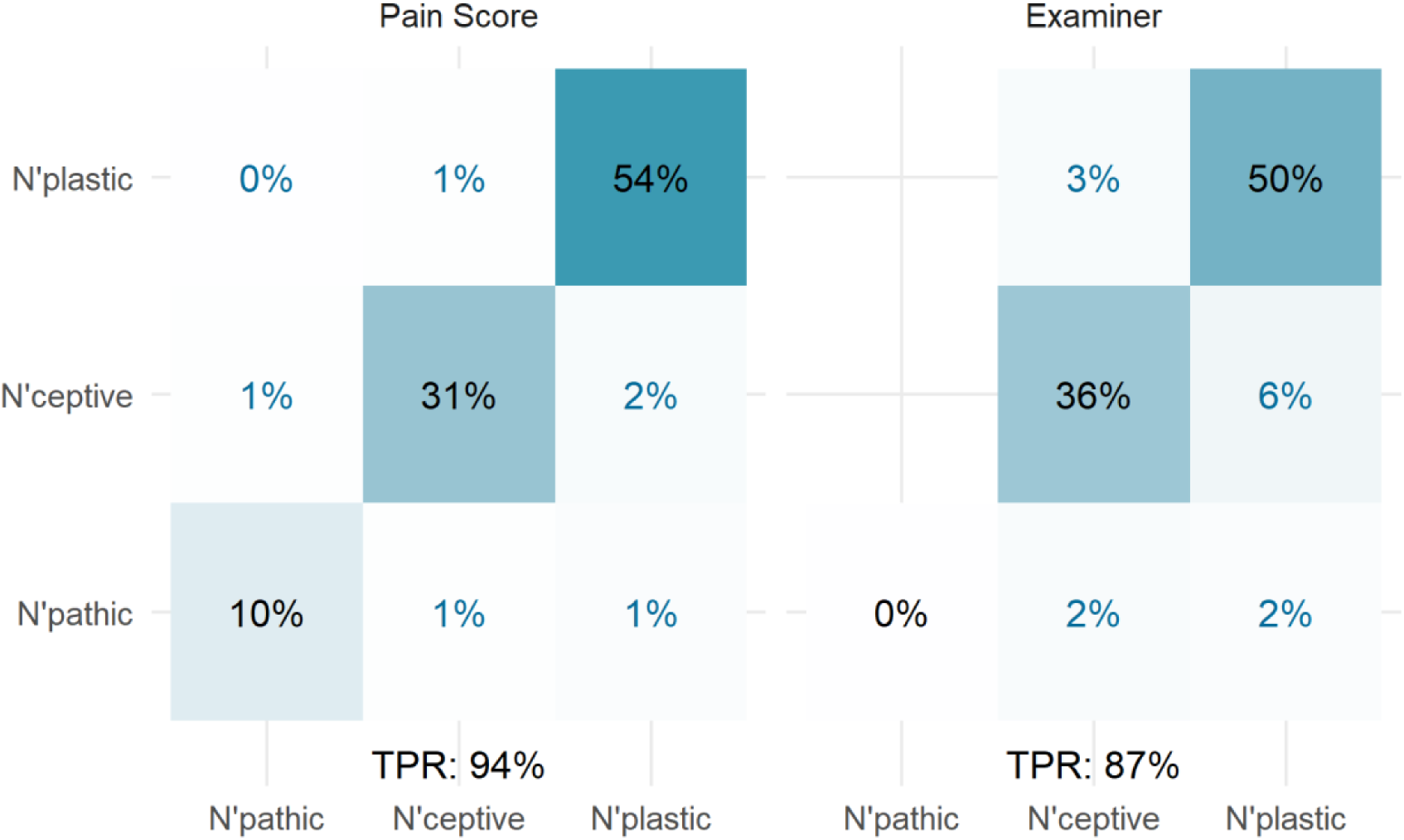
Supervised classification accuracy. Each of the pain type as computed with the score, and against the pain defined by the examiner were tested against a multiclass logistic regression model. Each comparison shows the percentage of predictions from one pain type that were classified as any pain type. The diagonal represents the correct predictions, and the sum of the diagonal is the True Positive Rate (TPR).

### Probabilistic predictions of pain mechanisms to evaluate coexistence of pain types

Both supervised and unsupervised machine learning techniques were used to generate probabilistic outcomes. This was used to address the complexity of pain type classification by giving insight into the coexistence of multiple pain types (that remains hidden in deterministic analysis). Figure 12 shows the comparison of 4 different probabilistic models, where the dominant cluster likelihood is represented as boxplots. The GMM have high concentration of values close to 100%, which is consistent with the FPC of 95%, the highest of the sample. In contrast, the supervised models (score-derived or examiner-derived pain types) have FPC in the range of 72-80%, and in both cases the neuropathic pain type presents the lowest likelihood values among all pain types. Supervised clustering highlights probability for coexistence of pain types, and greatest uncertainty for the neuropathic type.

**Figure 12.**
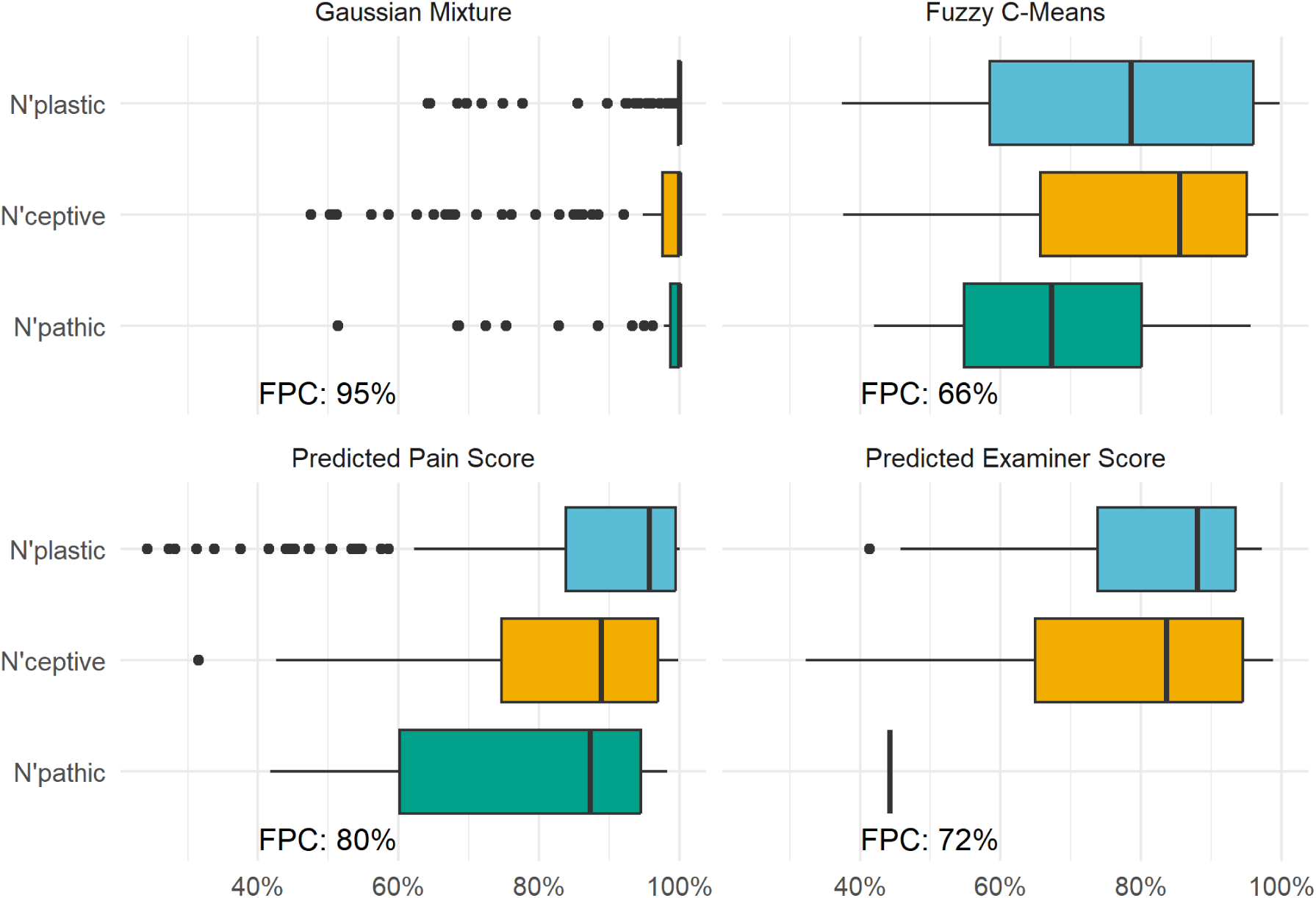
Probabilistic classification analysis. Each colour represents the dominant pain type, and the values represent the model likelihood for each participant. The top panels show the results of probabilistic clustering (unsupervised), and the lower panels represent the probabilistic predictions of the supervised models already analysed. Each model is accompanied by the Fuzzy Partition Coefficient (FPC) which measures the degree of uncertainty in the model predictions.

## Discussion

The results of this study support validity of clinical features to discriminate between pain types defined by IASP. Data were explained by three clusters; logistic regression demonstrated items that contribute substantially or little to the classification; and probabilistic analysis suggested some co-existence of pain types. This provides foundation to refine a tool to discriminate between pain types.

### Validity of features to discriminate between pain types

Without a gold standard method to assess pain types, our mathematical approach provided an innovative solution to investigate the validity of features to discriminate between them. Although this analysis cannot directly assess whether a participant’s pain was explained by the allocated type, three features support this interpretation – data were explained by three clusters, each cluster was characterised by high expression of features relevant for a specific pain type, and classification agreed with an experienced clinician (GMM - 82%). Allocation inaccuracy might be accounted for by mixed pain types (see below) or because criteria contributed equal weight to the cluster determination, yet the examiner might place more weight on specific features. Clinical criteria for nociplastic[15] and neuropathic[15] pain use weighting – e.g., subjective evidence of relevant pain distribution indicates *possible* neuropathic pain, whereas physical evidence carries greater weight as *probable* neuropathic pain. Lower TPR for other clustering methods relates to their properties. Unlike GMM which is a “soft” clustering technique enabling allocation of individuals to multiple clusters of different shapes[6], K-Means[18] and Hierarchical[32] clustering are “hard” techniques where individuals can only be allocated to a single cluster[13] (no mixed types), and clusters of different size or shape are problematic[21].

We considered pain types defined by the IASP[24], and identified in a systematic review[27]. Features used for classification are derived from items identified through a rigorous systematic review[28], expert consensus[29] and consideration of clinical criteria for neuropathic[5] and nociplastic[15] pain. Although it is possible that further pain types might have been identified if additional features were included, the current literature does not support this possibility.

### Relative importance of features

Supervised machine learning highlighted positive relationships for all features for each pain type, but with variation in coefficients. Coefficients from the examiner-derived pain type identify features that contributed substantially and little to classification (Table 2). This analysis could refine the features necessary to assess and requires consideration with clinical criteria for nociplastic and neuropathic pain.

For nociplastic pain important criteria were regional distribution, physical findings of mechanical/heat/cold sensitivity, and painful aftersensations. This aligns with IASP clinical criteria which consider regional pain to be “obligatory”. Evoked hypersensitivity phenomena (including painful aftersensations) are necessary for “possible” nociplastic pain and additional evidence of sensitivity is necessary for “probable” nociplastic pain[15]. No value was added by subjective evidence of sensitivity, which correlated with physical measures. Other “evoked hypersensitivity phenomenon” (physical/subjective evidence of sensitivity to light touch/allodynia) added little because of high correlation with aftersensations. Although comorbidities are commonly discussed as features of nociplastic pain[1,7] and prominent in the Central Sensitization Index[19], sensitivity to sound, etc, sleep disturbance, fatigue, and cognitive problems added little and they presented across pain types. Some argue comorbidities could be removed from diagnosis of nociplastic pain as they do not relate to nociceptive function[2].

For neuropathic pain, classification involved pain in a neuroanatomically plausible distribution, hypoaesthesia, and pain evoked by nerve provocation. No value was added by report of dysesthesia, lesion or disease to the somatosensory nervous system, or a neuropathic questionnaire score above cut-off pain. With reference to neuropathic pain clinical criteria[5], we did not formally test lesion or disease of somatosensory system which is required for “definite” neuropathic pain. We relied on patient report or available imaging. Physical evidence of pain and sensory symptoms within a neuroanatomically plausible distribution concur with “probable”. High correlation with subjective evaluation explains why physical measures added little. It is unsurprising that the Neuropathic Pain Questionnaire added little as this questionnaire predates the change in neuropathic pain definition from “dysfunction” to “lesion or disease” of the nervous system[12], and introduction of “nociplastic” pain[16]. The questions related to sensitivity would overlap between neuropathic and nociplastic pain[27].

There are no clinical criteria for nociceptive pain. The most important features included *exclusion* of other pain types. Features that should *not be* present were regional distribution (i.e., nociplastic), and hypoaesthesia (i.e., neuropathic). Features that should *be* present include no generalised hypersensitivity, which excludes nociplastic. Focus on exclusion of other pain types might be rectified by clinical criteria for nociceptive pain. The positive feature contributing to classification is pain provoked by specific postures/movements, which implies a response to tissue loading. This concurs with “movement evoked” or “mechanical” pain as alternative terms[27]. Unhelpful features include signs of inflammation and absence of autonomic signs which presented across pain types.

## Distribution and coexistence of pain types

The distribution of pain types is not generalisable. Participants were attending a chronic pain management program and might be biased to nociplastic pain. There will not yet be evidence of distribution of pain types because nociplastic pain was only recently introduced (although sharing features with “chronic primary pain”[22] or “central sensitization pain”[31] they are not synonymous), and methods are not available to discriminate between pain types (which motivated the present work).

Probabilistic analysis identified mixed pain types. An FPC of 100% indicates all patients allocated to a pain type could *only* be allocated to that pain type. This was not achieved for any pain type. Although consistent with reports of mixed pain types[27], it is not completely consistent with nociplastic clinical criteria which require “no evidence that nociceptive/neuropathic pain (a) is present or (b) if present, is entirely responsible for the pain”[15]. Although, this implies another mechanism can co-exist if it is not “entirely responsible” for the presentation, what that means is not clear. Identification of mixed pain types is important because this would enable tracking of changes in pain type over time – e.g., pain might begin as predominantly nociceptive pain, and transition to nociplastic features[14], and if classification of pain types is used to match patients to treatments[27], mixed pain types might impact their success.

Probabilistic analysis suggested greatest uncertainty for neuropathic pain, which could reflect the frequent presence of “sensitivity” in neuropathic pain[20,27,36]. A major motivator for “nociplastic” pain as the third pain type was the change in the definition of neuropathic pain from “dysfunction” to “disease or lesion” of nervous system pain[16]. It became necessary to account for patients who presented with sensitivity and symptoms unexplained by ongoing nociceptive input, and without nervous system damage or lesion[16]. Overlap of neuropathic and nociplastic could reflect some lack of clarity of this distinction.

### Development of a tool to discriminate between pain types

A tool is needed to discriminate between pain types, not only to provide insight into the plausible mechanisms underlying a patient’s pain, but also to match patients with treatments. This study applied a preliminary version of a tool, based on outcomes of expert consensus regarding features to support this decision[27]. This study provides initial validation of the features, and insight into features that are most important to inform classification.

Features of a tool to discriminate between pain types will differ from those for clinical criteria for a specific pain type because their purpose differs. As clinical criteria specify the features that must be present to confirm the presence of pain type and includes features that can be present in all pain types, they can confirm the presence of a pain type, but have limited capacity to exclude others or identify their co-existence. In contrast, as a tool designed to discriminate between pain types only uses features that are present in one (or two) pain types, it operationalises exclusion or co-existence of other pain types, but cannot confirm whether all criteria are met for a pain type. A hybrid tool will be required to satisfy both functions.

Further refinement of the pain type discrimination tool is required. First, features were judged clinically – operationalisation of these items would support interpretation and consistency. Second, weighting of features will be necessary. Third, the tool’s psychometric properties require evaluation. Fourth, evaluation of whether treatment outcomes are better when treatments are matched by the tool is important.

### Methodological limitations

There are several methodological considerations. First, features were scored using clinical judgement and we did not assess the psychometric properties. Second, we studied a diverse group with chronic musculoskeletal pain and might not be representative of other cohorts or environments. Third, features were based on a preliminary version of a tool and are likely to differ from final feature specification. Fourth, the experienced clinician used the clinical data to classify the pain types. Although initial cluster analysis did not use this information and is unbiased in its classification, these data were used for the supervised models with potential for bias. Fifth, additional clusters might be identified with additional features.

## Conclusion

In summary, this study provides initial validation of the approach to discriminate between pain types based on features of a patient’s pain presentation. This forms a foundation for future work to refine and a test tool to deploy in clinical practice.

## Data Availability

All data produced in the present work are contained in the manuscript

## Acknowledgments

Financial support was provided by the National Health and Medical Research Council of Australia (#2027473; #1194937). There are not relevant conflicts of interest.

---

**Suplementary Table 1.**
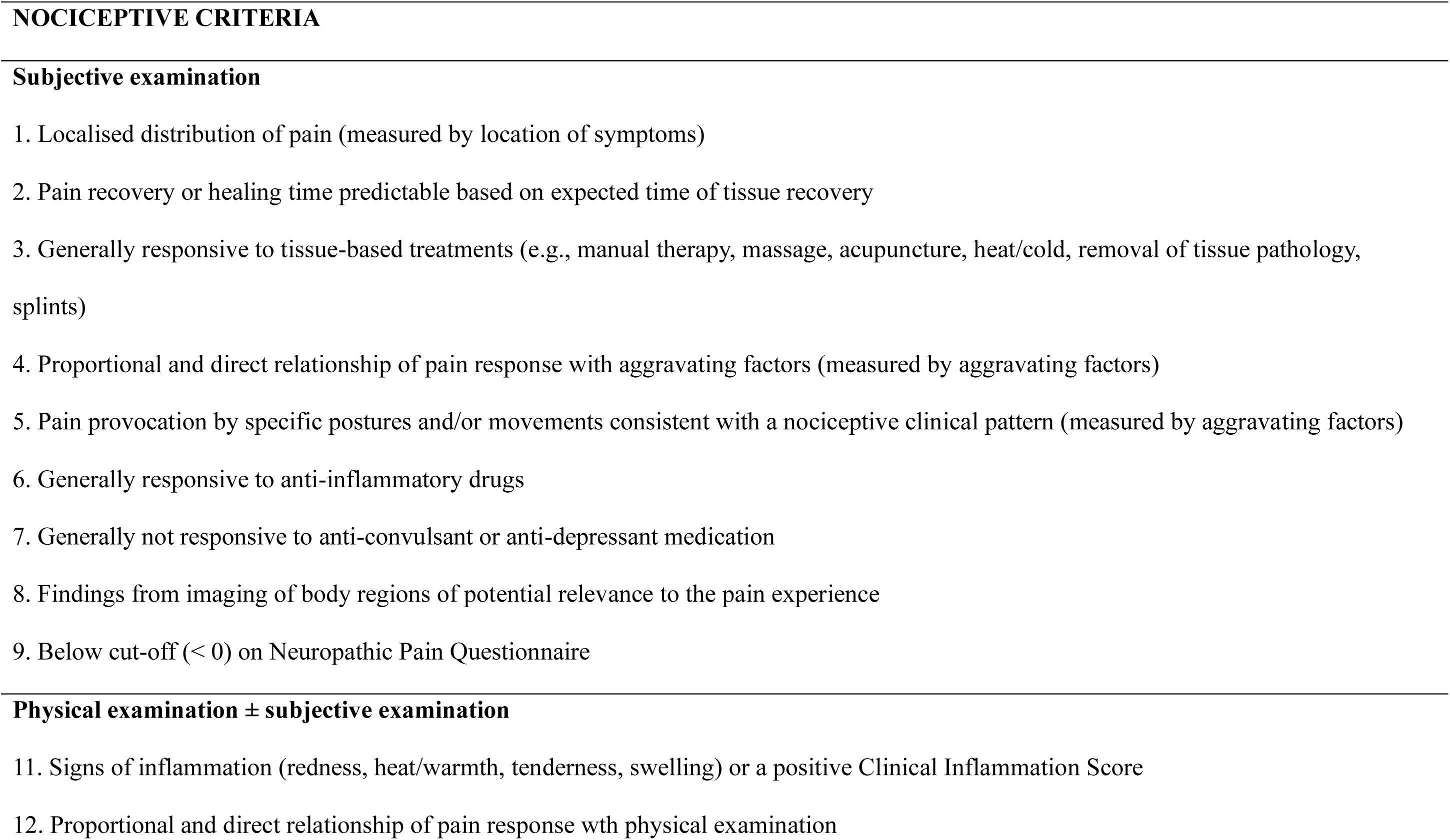
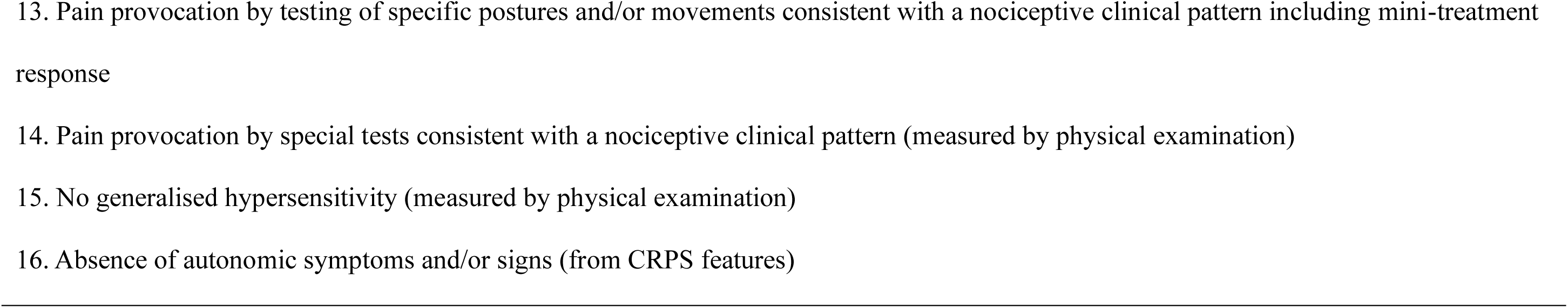
Clinical criteria for Nociceptive pain (based on [29])

**Supplementary Table 2.**
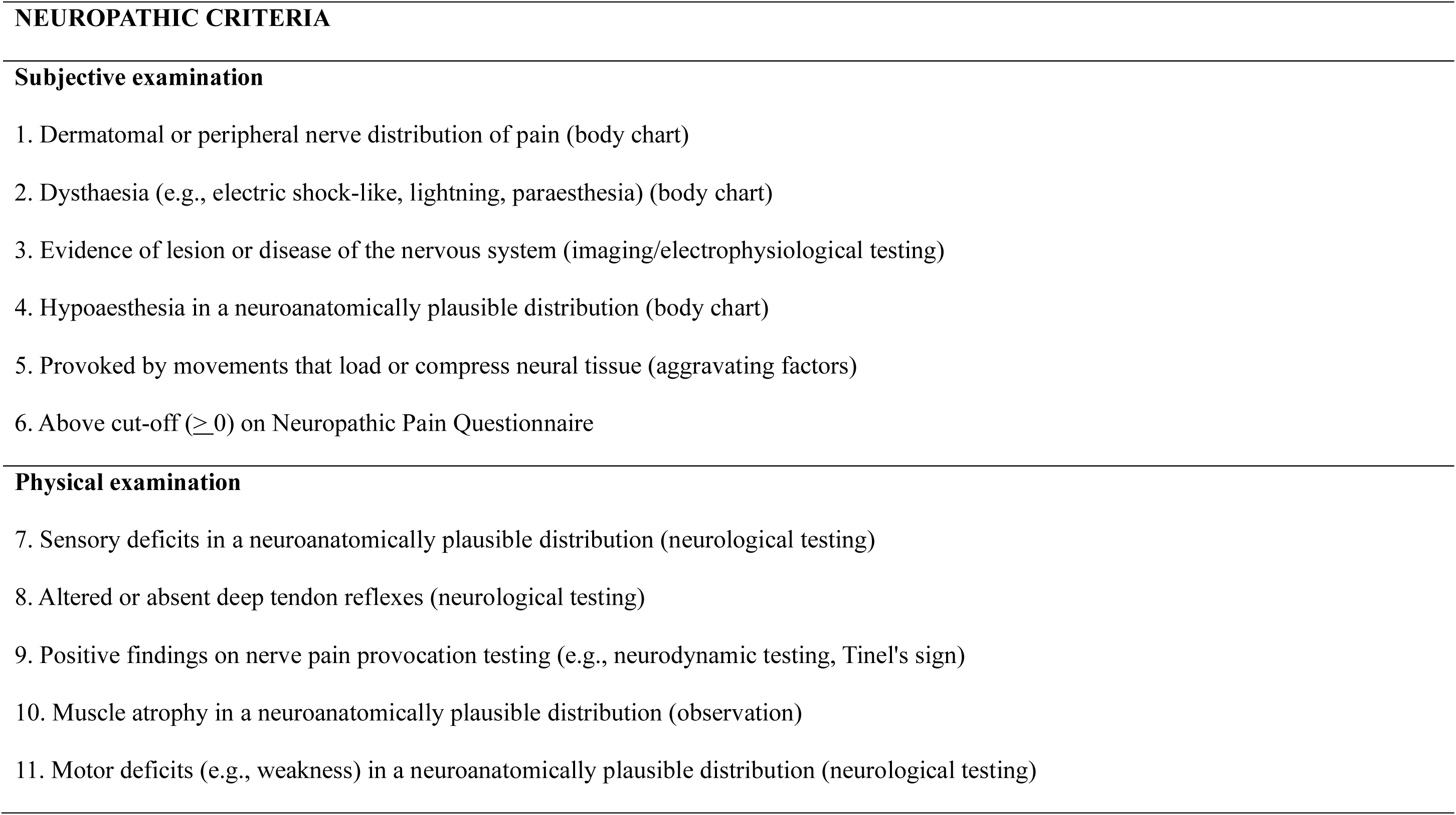
Clinical criteria for Neuropathic pain (based on [5])

**Supplementary Table 3.**
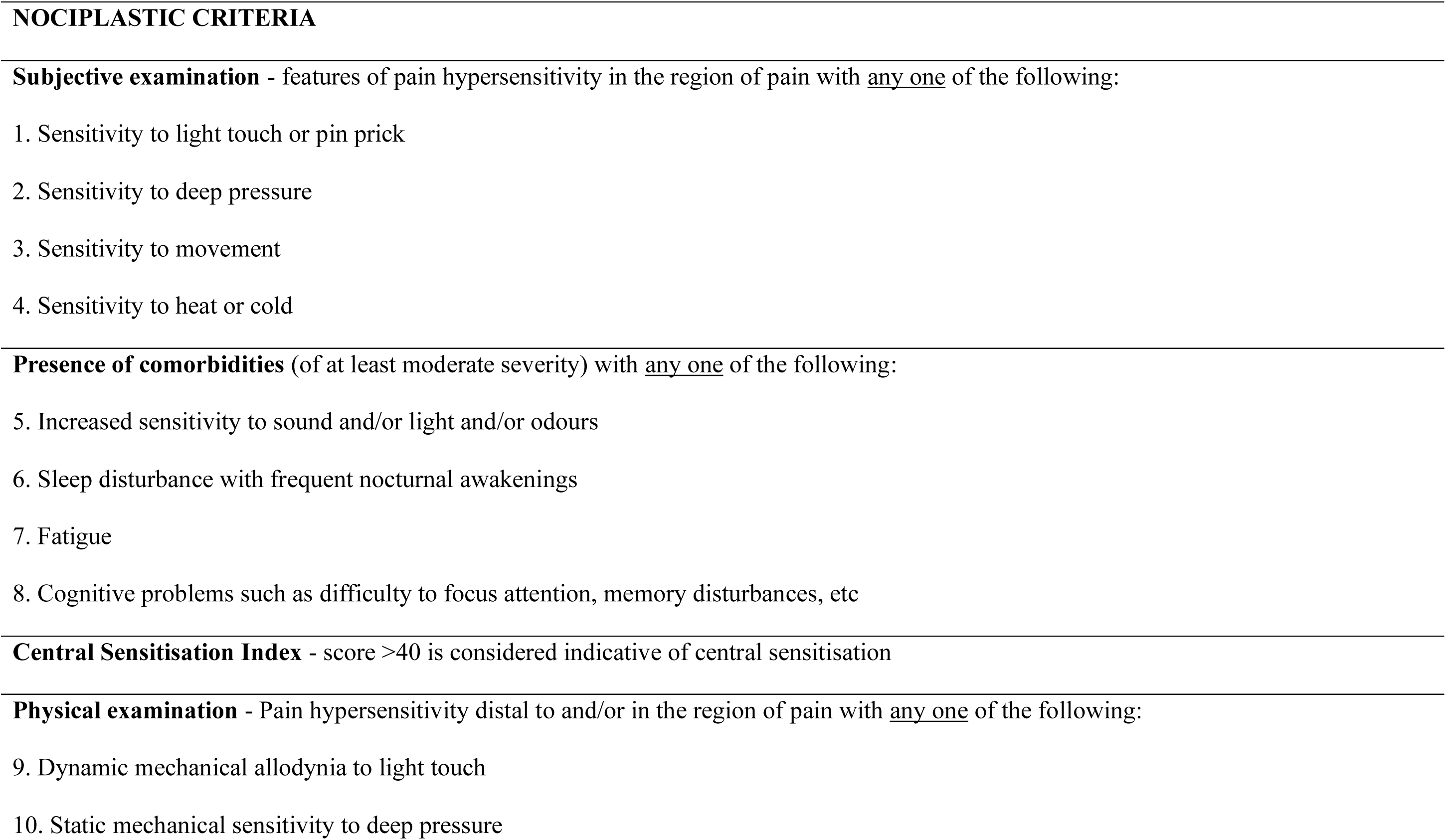
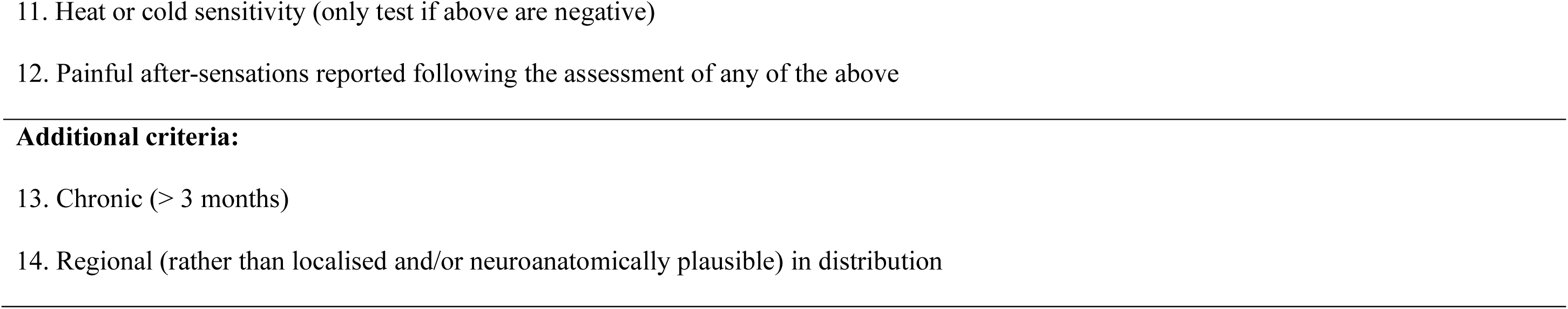
Clinical criteria for Nociplastic pain (based on [15])

